# COVID-19 Pandemic in Pakistan: Stages and Recommendations

**DOI:** 10.1101/2020.05.11.20098004

**Authors:** Farhan Saif

## Abstract

We present a real-time forecast of COVID-19 in Pakistan that is important for decision-making to control the spread of the pandemic in the country. The study helps to develop an accurate plan to eradicate the COVID-19 by taking calculated steps at the appropriate time, that are crucial in the absence of a tested medicine. We use four phenomenological mathematical models, namely Discrete Exponential Growth model, the Discrete Generalized Growth model, the Discrete Generalized Logistic Growth, and Discrete Generalize Richards Growth model. Our analysis explains the important characteristics quantitatively. The study leads to understand COVID-19 pandemic in Pakistan in three evolutionary stages, and provides understanding to control its spread in the short time domain and in the long term domain. For the reason the study is helpful in devising the measures to handle the emerging threat of similar outbreaks in other countries.

## 1 Introduction

The COVID-19 causes severe acute respiratory syndrome. It have been linked to a live animal seafood market in Wuhan, pointing to a zoonotic origin of the epidemic. Human-to-human transmission, however, has driven its spread in the strongly interconnected human world community [1, 2]. While the transmission potential of novel coronavirus can reach high values the epidemiological features of COVID-19 are still unclear [3]. On December 27, 2019, a hospital in Wuhan, capital of Hubei province, reported the first mysterious, suspected pneumonia cases to the center of disease control (CDC) in the capital. On January 8, 2020 a new coronavirus was identified as the cause of pneumonia that spread all over China in next three weeks. Since then, the novel coronavirus, COVID-19, spread across the globe in the next month. In March the World Health Organization declared it to be a pandemic, that in the absence of a vaccine became a challenge for mankind with a tendency to grow exponentially if no measure is taken to prevent it [4]. The incubation period of COVID-19 can last for two weeks or longer [5]. During the period of latent infection, the disease may still be infectious [6]. The virus can spread from person to person through respiratory droplets and close contact [7].

The government of Pakistan has taken a number of measures, including the implementation of hygienic awareness, isolation and quarantining the patients, closure of important entry points on international borders, closure of educational institutions, parks, business centres, postponement of national and international flights, the launch of clinical trials, and epidemiological surveillance, with the aim of protecting the population. Together with these efforts an optimal allocation of prevention measures, medical resources, organization of production activities, and, eventually, maintenance of national economic development throughout the country require calculated steps. In the present contribution we develop analytical approach based on phenomenological models, namely Discrete Exponential Growth model, the Discrete Generalized Growth model (DG^2^M), the Discrete Generalized Logistic Growth (DGLM), and Discrete Generalize Richards Growth model (DGRM). The analytical work makes it possible to make useful prediction in the short time domain and in the long time domain to control the exponentially growing pandemic in the country.

The epidemic study is performed by various mathematical models which include SIR model [8], SEIR model [9], generalized SEIR model [10] and neural network based quarantine control model [11]. However, the mathematical analysis of pandemics based on exponential growth model [12, 13, 14, 15] and generalized growth model [16, 17, 18, 19, 20, 21, 22] have been validated in previous outbreaks, such as Ebola virus, influenza, smallpox, plague, measles, HIV/AIDS, severe acute respiratory syndrome, Zika virus, COVID-19 and identified as the best model to adequately fit the data [25, 26, 27, 28, ?]. The Discrete Exponential Growth model, Discrete Generalized Growth model (DG^2^M), Discrete Generalized Logistic model (DGLM), and Discrete Generalized Richard model (DGRM) measure characteristics values including the growth in the outbreak, rate of increase and the size of the outbreak, and evolution trend that leads to limit the pandemic. Moreover, these models perform the microscopic and macroscopic study of pandemics. Based on our numerical simulations we find excellent agreement between the analytical results and the real-time data of coronavirus in Pakistan.

The manuscript has the following layout: in section 2 we present the mathematical model for our study, in section 3 we explain the real-time data of confirmed COVID-19 cases and present evolutionary stages of the pandemic in Pakistan. In section 4 we present our conclusions and recommendations.

## 2 Mathematical Model

The Exponential Growth model, the Generalized Growth model, the Generalized Logistic Growth, and Generalized Richards Growth model [30], are expressed by the differential equations,

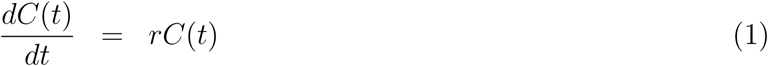

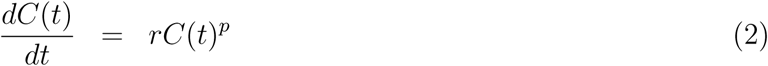

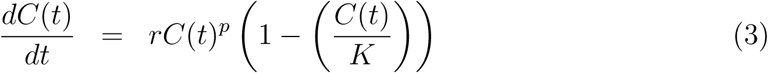

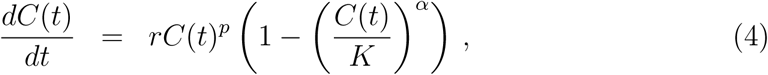

respectively. These have been considered extensively to explain previous epidemics. Here C(*t*) describes the cumulative number of cases at time *t*. The equation (4) is the extension of the original Richards growth model [31] which explain the pandemics [32, 33, 34]. Here, *r* is the growth rate at the early stage, and *K* is the final epidemic size. Moreover *p* is a parameter that allows the model to capture different growth profiles. The exponent *α* measures the deviation from the dynamics of the simple logistic curve. The equation (4) corresponds to the original Richards model for *p* = 1, that is

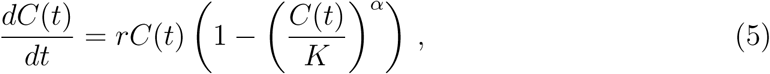

and reduces to the generalized logistic model for *α* =1 and *p* =1.

Keeping in view the development of data in terms of the positive tests, variations, and deaths on every next day we may consider the change in *C* over a unit interval of time. We express the rate of change of *C*, that is 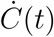, in difference form as 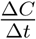.

Here Δ*C* = *C_n_*_+1_ − *C_n_* is the change in positively tested patients. Thus we reshape the equation (4) in the discrete form as

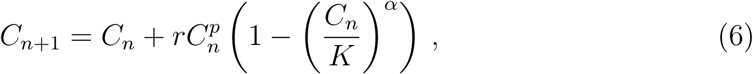

where *C_n_* is the cumulative number of cases on the *n*th day. The discrete generalized Richard model (DGRM) provides an effective way to monitor the pandemic in the form of a mapping, thus connecting the data on *n*th day with (*n* + 1)th day. The stable points of the discrete generalized Richards model given in equation (6) are *C_n_* = 0, that implies no positively tested patients, and *C_n_* = *K*. The stable points are the same as obtained by the generalized Richards model.

We write the discrete generalized growth model (DG^2^M), as

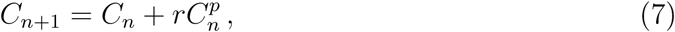

which provides exponential growth for *p* = 1, that is 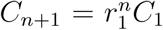, that relates the number of positively tested patients on the first day with the number of postively tested patients after a number of *n* days has passed, here *r*_1_ = 1 + *r*. Hence for the parameter *p* = 0 we find linear increase with respect to n as *C_n_*_+1_ = *C*_1_ + *nr*, and sub-exponential growth for 0 < *p* < 1.

## 3 Evolutionary Stages of COVID19 in Pakistan

To control the COVID-19 pandemic in the absence of medicines, the requirement is to take calculated steps at the appropriate time. Therefore in addition to data analysis, we need a visible understanding, that is necessary for the management of the associated risk to the public life and economy. The day-to-day development of the pandemic in all the provinces of Pakistan describes an exponential increase of the confirmed COVID-19 cases that include active, recovered, and deaths (expressed by blue dots in figure 1) that is verified mathematically using discrete generalized growth model (DG^2^M), discrete logistics growth models (DGM) and discrete generalized Richard model (DGRM). The models have been used to comprehend the COVID-19 outbreak in China, USA, Europe and the other countries of the world [36].

**Figure 1:**
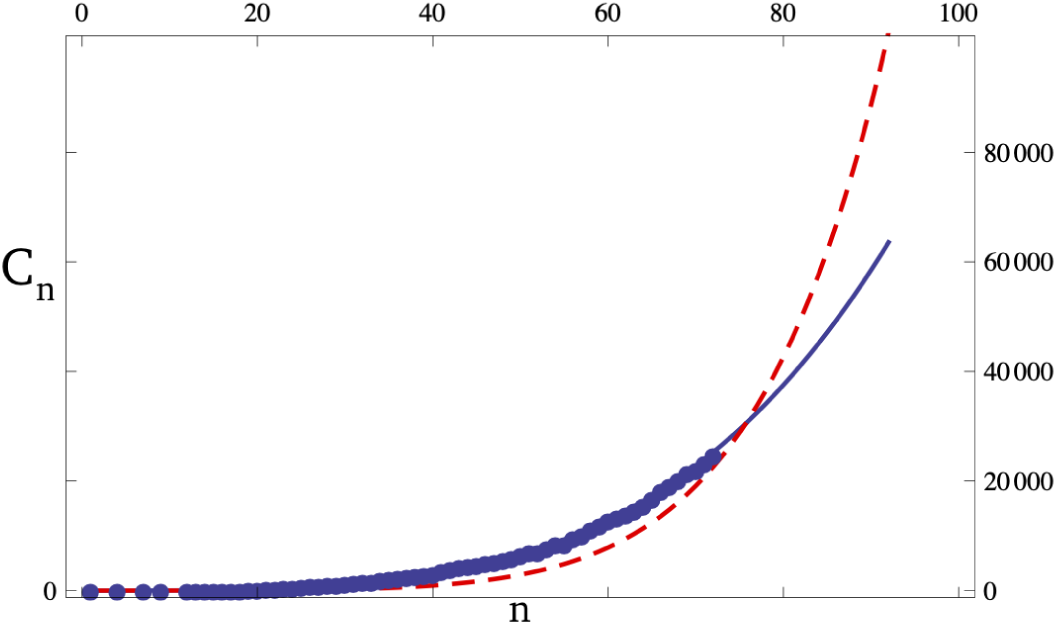
Evolution of confirmed COVID-19 patients (*C_n_*) in Pakistan plotted versus number of days, *n*. The real time data is presented by blue dots. The number of confirmed cases in the first twenty days follows DG^2^M curve for *r* = 0.3 and *p* = 0.875 (red dashed line). The real time data in stage III follows DG^2^M curve for *r* = 0.74 and *p* = 0.74 (blue line).

### Stage I - February 26 to March 15

On February 26, 2020 first two confirmed coronavirus cases were reported in Pakistan. In the absence of lockdown and public awareness campaign in the country, in the next two weeks the pandemic spread exponentially all over the country. The comparison of the real-time data with the numerical results based on DG^2^M model endorses the exponential growth of the pandemic with an exponent value 0.85. (The DG^2^M result is shown by the red dashed line in figure 1 and 2.)

**Figure 2:**
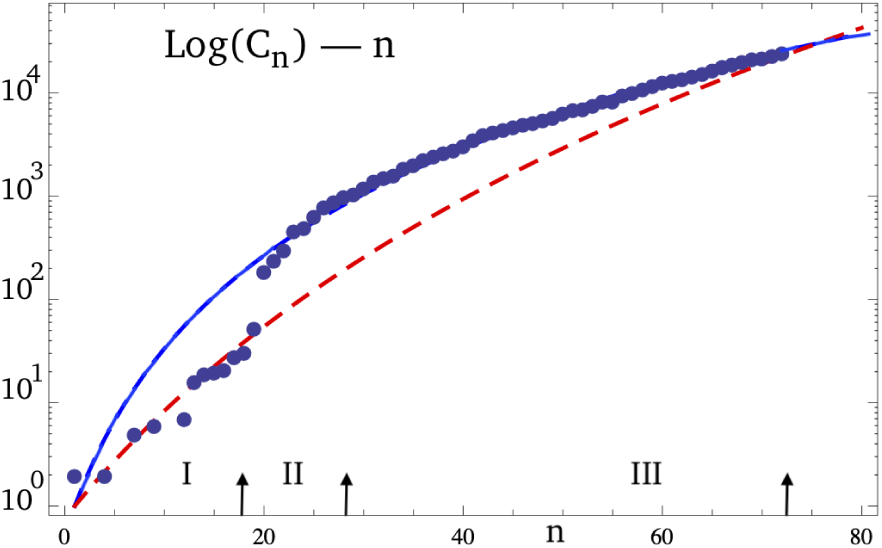
In the log-plot number of confirmed COVID-19 cases in Pakistan, *log*(*C_n_* are plotted versus the number of days *n*, from February 26, 2020. The real time data is presented by blue dots. The number of confirmed cases in the first twenty days follows DG^2^M curve for *r* = 0.3 and *p* = 0.875 (red dashed line). The real time data follows DG^2^M curve for *r* = 0.74 and *p* = 0.74 (blue line).

### Stage II- March 16 to March 31

The arrival of zaireen from Iran, participants in tableeghi jamat from foreign countries, and Pakistani people arriving from abroad in special flights caused an abrupt increase in the COVID 19 confirmed cases in the fifteen days, from March 16 to March 31 (seen in the figure 2). The unexpected burden of thousands of people with foreign travel history, in the presence of lack of testing and quarantine facilities, and transporting the zaireen to their home towns caused enormous pressure on limited financial resources and medical infrastructure. The poor state of management pushed the country many weeks ahead in time in its fight against growing COVID-19 pandemic.

### Stage III: April 01- May 08

In the next thirty eight days de-facto quarantining of the provinces by restricting the movement of people, employing social distancing, tracing and tracking, closing educational institutes, parks, and restaurants, limiting air and train travel services, the Government of Pakistan tried to control the COVID-19 spread in the country. This reduced the exponential growth from 0.85 (the stage I value) to the new exponent value as 0.74 (the DG^2^M result is shown by blue line in figure 2).

The measures to eradicate COVID-19 pandemic in Pakistan during stage III resulted in the number of confirmed cases today as 28736, including 20291 active cases, 7809 recovered and 636 deaths. The mathematical analysis displays an intersection of red and blue dashed lines, that corresponds to second week of May in real-time. This implies that the number of confirmed COVID-19 cases were the same today in the absence of any lockdown and public awareness campaign at stage I, if in stage II the country had allowed the people with foreign travel history to enter only after adequate check up and compulsory quarantine.

All the measures to ensure lockdown caused a blow to economy of the country at stage II and III. Eventually they appeared to be a cost of the mismanagement in handling the people entry with a foreign travel history. At the same time it displayed the unprepared state to handle the pandemic in Pakistan at the early stages.

### Stage IV: May 09- May 31

The analysis of the COVID-19 confirmed cases based on DGRM and its extrapolation to May 31 show that with presently adopted lockdown the pandemic size shall increase to around sixty thousands confirmed cases in the next twenty two days (shown in figure 3, narrow-dashed black line). It further expresses that by intensifying lockdown at this stage we can appreciably slow down the pandemic in Pakistan by end of June, 2020. Thus instead of relaxing the lock-down, a strict lockdown is useful at this stage, therefore enabling Pakistan to control the pandemic in next two months. This way the size of the outbreak is restricted to around one hundred thousands confirmed cases, as seen in figure 4.

**Figure 3:**
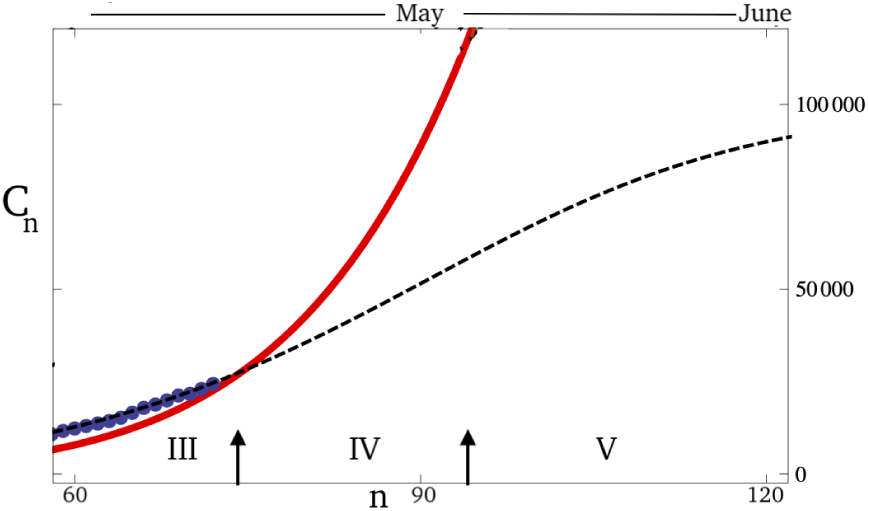
Evolution of confirmed COVID-19 patients (*C_n_*) in Pakistan plotted versus number of days, *n*. The real time data is presented by blue dots, the DG^2^M result is plotted (by red line) for *r* = 0.3 and *p* = 0.875. DGRM results is plotted (in black dashed line) for *r* = 0.74 and *p* = 0.74, *α* = = 2 and K = 100000.

**Figure 4:**
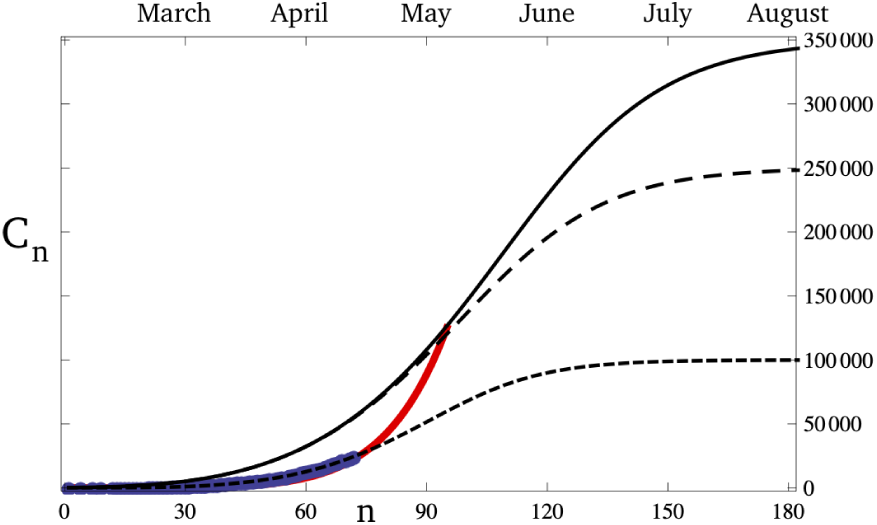
Evolution of Confirmed COVID-19 patients (*C_n_*) plotted versus number of days, *n*. The real time data is presented by blue dots, the DG^2^M result is plotted (by red line) for *r* = 0.3 and *p* = 0.875. DGRM results is plotted (in black dashed line) for *r* = 0.74 and *p* = 0.74, *α* = 2 and K = 100000. DGRM results plotted in black big-dashed line and black line for K = 250000 and K = 350000.

In case the lockdown disappears completely at stage IV the pandemic size would increase to one hundred twenty thousand confirmed COVID-19 cases in next twenty two days, as indicated by the red line in figure 3. Lifting the lockdown partially shall bring the numbers of confirmed COVID-19 cases in between sixty thousand and one hundred twenty thousand.

### Stage V: June 01-

Keeping in view the month of Ramadan and the necessary business requirements in these days, the devised lockdown in stage II and III is relaxed in the stage IV. The pandemic management in next months requires a careful and calculated approach.

No lockdown scenario from May 09 to May 31 shall drag the size of pandemic in the country to higher values. By extrapolating the DGRM results we note that a strict lockdown in June shall bring the number of confirmed cases in September around three hundred fifty thousands, with less than one percent increase in new cases per day, as shown by curved black line in figure 4. The peak is expected to be at the end of June with around two hundred thousands confirmed cases.

A compromised lockdown at stage IV shall make it possible to control the COVID-19 pandemic by end of July, with the estimated size of the outbreak around two hundred fifty thousand confirmed cases, as shown by big-dashed black line in figure 4.

## 4 Conclusions

In order to avoid business melt down and economic depression in Pakistan the Government is easing the lockdown measures from May 09 till May 31. The business centers, shops, and train travel services are opening in these days, anticipating business activities and people transportation. The within-a-city and inter-city movement of people shall make the public vulnerable and take the COVID-19 pandemic to higher levels.

We show that by easing the lockdown measures in Pakistan the COVID-19 pandemic may increase four time more than the present value in next three weeks. Therefore with the given relaxation of lockdown in the month of May, a parallel public service campaign is to be launched by the government of Pakistan. This way people must be made concious of the growing pandemic situation in Pakistan all the time. The doctors, subject experts, university faculty, educationalists, and media should play an active role to keep the people well aware of the seriousness of the pandemic and convince them to take every measures essential for their own security.

The future risk management requires a stringent lockdown in June to control the pandemic in the country. The effective preparedness on medical, and technical fronts together with optimal allocation of prevention measures, resources, and organization of production activities shall take the country out of the global COVID-19 pandemic. Mismanagement and delayed actions however have tendency of increasing in the size of pandemic and economical loss in next months.

## Data Availability

The data is collected from the Government of Pakistan official page and Wikipedia

## References

[1] Roosa K, Lee Y, Luo R, Kirpich A, Rothenberg R, Hyman JM, Yan P, Chowell G. 2020a. Real-time forecasts of the COVID-19 epidemic in China from February 5th to February 24th, 2020. Infect Disease Modell, 5: 256e263. DOI: 10.1016/j.idm.2020.02.002.

[2] Bianconi A, Marcelli A, Campi G, Perali A. 2020. Ostwald growth rate in controlled Covid-19 epidemic spreading as in arrested growth in quantum complex matter. arXiv:2003.08868.

[3] Roosa K, Lee Y, Luo R, Kirpich A, Rothenberg R, Hyman JM, Yan P, Chowell G. 2020b. Short-term forecasts of the COVID-19 epidemic in Guangdong and Zhejiang, China: February 13–23, 2020. J Clin Med, 9: 596, DOI:10.3390/jcm9020596.

[4] Joint Mission WHO. Report of the who-china joint mission on coronavirus disease 2019 (covid-19), 2020. URL https://www.who.int/docs/default-source/coronaviruse/who-china-joint-mission-on-covid-19-final-report.pdf. (accessed March 28, 2020).

[5] World Health Organization, “Report of the WHO-China Joint Mission on Coronavirus Disease 2019 (COVID-19),” World Health Organization, 2020.

[6] Ruiyun Li, Sen Pei, Bin Chen, Yimeng Song, Tao Zhang, Wan Yang, and Jeffrey Shaman. Substantial undocu-mented infection facilitates the rapid dissemination of novel coronavirus (sars-cov2). Science, 2020.

[7] Coronavirus Disease 2019 (COVID-19), “Centers for Disease Control and Prevention, 2020. [Online]. Available: https://www.cdc.gov/coronavirus/2019-ncov/about/symptoms.html. [Accessed 27 February 2020].

[8] Waqas M., Farooq M., Ahmad R., and Ahmad A., Analysis and Predictions of COVID 19 pandemic in Pakistan using time dependent SIR model, arXiv: 2005.02353v1

[9] Binti Hamzah FA, Lau C, Nazri H, Ligot DV, Lee G, Tan CL, et al. CoronaTracker: World-wide COVID-19 Outbreak Data Analysis and Prediction. [Submitted]. Bull World Health Organ. E-pub: 19 March 2020. doi: http://dx.doi.org/10.2471/BLT.20.255695

[10] Lenka Pribiylova and Veronika Hajnova, SEIAR model with asymptomatic cohort and consequences to efficiency of quarantine government measures in COVID-19 outbreak, arXiv:2004.02601-v1

[11] Raj Dandekar and George Barbastathis, Neural Network aided quarantine control model estimation of global Covid-19 spread, arXiv:2004.02752v1

[12] Tuite AR, Fisman DN, Reporting Epidemic Growth and Reproduction Numbers for the 2019 Novel Coronavirus (2019-nCoV) Epidemic. Ann Intern Med. 2020 (February):2019–20

[13] Zhao S, Cao P, Gao D, Zhuang Z, Chong MKC, Cai Y. Epidemic growth and reproduction number for the novel coronavirus disease (COVID-19) outbreak on the Diamond Princess cruise ship from January 20 to February 19, 2020: A preliminary data-driven analysis. SSRN. 2020. Preprint at: https://ssrn.com/abstract=3543150

[14] You C, Deng Y, Hu W, Sun J, Lin Q, Zhou F, et al. Estimation of the Time-Varying Reproduction Number of COVID-19 Outbreak in China.

[15] Zhang S, Diao M, Yu W, Pei L, Lin Z, Chen D. Estimation of the reproductive number of Novel Coronavirus (COVID-19) and the probable outbreak size on the Diamond Princess cruise ship: A data-driven analysis. Int J Infect Dis. 2020.

[16] Li Y, Yin X, Liang M, Liu X, Hao M, Wang Y. A Note on NCP Diagnosis Number Prediction Model. medRxiv. 2020. Preprint at: https://www.medrxiv.org/content/10.1101/2020.02.19.20025262v1

[17] Maier BF, Brockmann D. Effective containment explains sub-exponential growth in confirmed cases of recent COVID-19 outbreak in Mainland China. arXiv. 2020. Preprint at: https://arxiv.org/abs/2002.07572

[18] Ying S, Li F, Geng X, Li Z, Du X, Chen H, et al. Spread and control of COVID-19 in China and their associations with population movement, public health emergency measures, and medical resources. medRxiv. 2020. Preprint at: https://www.medrxiv.org/content/10.1101/2020.02.24.20027623v1

[19] Brandenburg A. Quadratic growth during the 2019 novel coronavirus epidemic. arXiv. 2020. Preprint at: http://arxiv.org/abs/2002.03638

[20] Ziff AL, Ziff RM. Fractal kinetics of COVID-19 pandemic. medRxiv. 2020. Preprint at: https://www.medrxiv.org/content/10.1101/2020.02.16.20023820v2

[21] Muniz-Rodriguez K, Chowell G, Cheung C-H, Jia D, Lai P-Y, Lee Y, et al. Epidemic doubling time of the COVID-19 epidemic by Chinese province. medRxiv. 2020. Preprint at: https://www.medrxiv.org/content/10.1101/2020.02.05.20020750v4

[22] Zhang J, Litvinova M, Wang W, Wang Y, Deng X, Chen X, et al. Evolving epidemiology of novel coronavirus diseases 2019 and possible interruption of local transmission outside Hubei Province in China: a descriptive and modeling study. medRxiv. 2020. Preprint at: https://www.medrxiv.org/content/10.1101/2020.02.21.20026328v1

[23] Roosa K, Lee Y, Luo R, Kirpich A, Rothenberg R, Hyman JM, et al. Short-term Forecasts of the COVID-19 Epidemic in Guangdong and Zhejiang, China: February 13–23, 2020. J Clin Med. 2020.9(2):596 abstract id=3539658

[24] Lin H, Liu W, Gao H, Nie J, Fan Q. Trends in Transmissibility of 2019 Novel Coronavirus-infected Pneumonia in Wuhan and 29 Provinces in China. SSRN. 2020. Preprint at: https://papers.ssrn.com/sol3/papers.cfm7abstractid=3544821

[25] Viboud C, Simonsen L, Chowell G. A generalized-growth model to characterize the early ascending phase of infectious disease outbreaks. Epidemics. 2016;15:27–37.

[26] Chowell G, Tariq A, Hyman JM. A novel sub-epidemic modeling framework for short-term forecasting epidemic waves. BMC Med. 2019;17:164. doi:10.1186/s12916-019-1406-6.

[27] Wu K, Darcet D, Wang Q, Sornette D. Generalized logistic growth modeling of the COVID-19 outbreak in 29 provinces in China and in the rest of the world. 2020. http://arxiv.org/abs/2003.05681. Accessed 4 Apr 2020.

[28] Koczkodaj WW, Mansournia MA, Pedrycz W, Wolny-Dominiak A, Zabrod-skii PF, Strzaška D, et al. 1,000,000 cases of COVID-19 outside of China: The date predicted by a simple heuristic. Glob Epidemiol. 2020;:100023. doi:10.1016/j.gloepi.2020.100023.

[29] Rouamba T., Samadoulougou S., Bonnechère B., Chiem B. and Kirakoya-Samadoulougou F., What Can We Learn from Burkina Faso COVID-19 Data? Using Phenomenological Models to Characterize the Initial Growth Dynamic of the Outbreak and to Generate Short-Term Forecasts, doi:10.20944/preprints202004.0328.v1

[30] Ke Wu and Didier Darcet and Qian Wang and Didier Sornette, Generalized logistic growth modeling of the COVID-19 outbreak in 29 provinces in China and in the rest of the world, arXiv:2003.05681.

[31] Richards FJ. A flexible growth function for empirical use. J Exp Bot. 1959.10(2):290–301.

[32] Chowell G. Fitting dynamic models to epidemic outbreaks with quantified uncertainty: A primer for parameter uncertainty, identifiability, and forecasts. Infect Dis Model. 2017.2(3):379–98.

[33] Chowell G, Hincapie-Palacio D, Ospina J, Pell B, Tariq A, Dahal S, et al. Using Phenomenological Models to Characterize Transmissibility and Forecast Patterns and Final Burden of Zika Epidemics. PLoS Curr. 2016.8.

[34] Chowell G, Luo R, Sun K, Roosa K, Tariq A, Viboud C. Real-time forecasting of epidemic trajectories using computational dynamic ensembles. Epidemics. 2020.30(November 2019):100379.

[35] https://www.populationpyramid.net/united-states-of-america/2019/

[36] Saif F., Signature of the State measures on the COVID-19 Pandemic in China, Italy, and USA, https://doi.org/10.1101/2020.04.08.20057489.

[37] Peter Forster, Lucy Forster, Colin Renfrew, and Michael Forster, Phylogenetic network analysis of SARS-CoV-2 genomes, PNAS first published April 8, 2020 https://doi.org/10.1073/pnas.2004999117

